# Modeling Influenza Seasonality in the Tropics and Subtropics

**DOI:** 10.1101/2021.02.04.21251148

**Authors:** Haokun Yuan, Sarah C. Kramer, Eric H. Y. Lau, Benjamin J. Cowling, Wan Yang

## Abstract

Climate drivers such as humidity and temperature may play a key role in influenza seasonal transmission dynamics. Such a relationship has been well defined for temperate regions. However, to date no models capable of capturing the diverse seasonal pattern in tropical and subtropical climates exist. In addition, multiple influenza viruses could cocirculate and shape epidemic dynamics. Here we construct seven mechanistic epidemic models to test the effect of two major climate drivers (humidity and temperature) and multi-strain co-circulation on influenza transmission in Hong Kong, an influenza epidemic center located in the subtropics. Based on model fit to long-term influenza surveillance data from 1998 to 2018, we found that a simple model incorporating the effect of both humidity and temperature best recreated the influenza epidemic patterns observed in Hong Kong. The model quantifies a bimodal effect of absolute humidity on influenza transmission where both low and very high humidity levels facilitate transmission quadratically; the model also quantifies the monotonic but nonlinear relationship with temperature. In addition, model results suggest that, at the population level, a shorter immunity period can approximate the co-circulation of influenza virus (sub)types. The basic reproductive number *R*_*0*_ estimated by the best-fit model is also consistent with laboratory influenza survival and transmission studies under various combinations of humidity and temperature levels. Overall, our study has developed a simple mechanistic model capable of quantifying the impact of climate drivers on influenza transmission in (sub)tropical regions. This model can be applied to improve influenza forecasting in the (sub)tropics in the future.

## Introduction

Influenza is a disease of considerable public health concern, causing roughly 300,000-650,000 deaths and 3-5 million cases of severe illness each year worldwide [1]. Although evidence suggests that the burden of influenza in the tropics and subtropics is not substantially less than in temperate regions [2, 3], studies on influenza in these regions are comparatively rare. Likewise, modeling and forecasting efforts, which may promote both understanding of and preparation for outbreaks in the future, have mostly been focused on countries with temperate climates [4, 5]. A variety of factors contribute to this disparity, including lack of long-term surveillance data and competing public health interests [6-8]. We focus here on two features of influenza epidemics in these regions that particularly complicate modeling efforts, i.e., the lack of understanding of climatic drivers and cocirculation of multiple influenza types and subtypes.

In temperate regions, influenza displays a clear seasonal pattern, with epidemics occurring in the winter and very few cases observed during the summer [9, 10]. While several potential drivers of this pattern have been suggested [9], humidity appears to be particularly important in driving these “seasonal” influenza epidemics. Specifically, both survival and transmission of the influenza virus are heightened when absolute humidity (AH) is low [11], corresponding to the yearly observed peaks of influenza activity in winter [12].

In tropical or subtropical locations, this seasonal pattern is less commonly observed. Instead, influenza causes multiple epidemics each year, or else is present year-round with unpredictable variation in intensity [3, 9, 10, 13]. In addition, humidity is relatively high all year in the tropics and subtropics, and influenza epidemics tend to occur during the rainy season, when humidity is particularly high [13, 14]. Thus, the relationship observed in temperate regions and modeled by Shaman et al. [12], where influenza transmission decreases monotonically with increasing AH, is not sufficient to explain patterns in influenza transmission in the tropics and subtropics [9, 15].

While the exact impacts remain unclear, humidity, precipitation and temperature are the main contenders as climate drivers for influenza transmission in the tropics and subtropics. A few studies suggest that the impact of humidity on influenza transmission may be bimodal, rather than unimodally decreasing as suggested by Shaman at al. [11, 12]. Work by Yang et al. showed that influenza virus survival is higher at lower relative humidity (<50%), but also found increased survival at very high levels of relative humidity (∼100%) [16]. By analyzing patterns of influenza transmission in 78 locations worldwide, Tamerius et al. [14] found that influenza outbreaks in locations that experience high temperature and high AH year-round had a tendency to occur during the rainy season, when both AH and rainfall were high. This pattern has been consistently reported in several countries [6, 17-19]. Deyle et al. [20] instead suggested that influenza transmission is driven by AH, moderated by temperature. Specifically, they find that influenza activity decreases with increasing AH up to about 24°C (75 °F), then increases with increasing AH up to about 30°C (86°F), at which point high temperatures strongly restrict influenza transmission.

In addition to diverse climate drivers, co-circulation of multiple influenza viruses may contribute to the observed multiple yearly influenza epidemics in the tropics and subtropics. Circulating human influenza viruses are classified into types (A or B, based on genus) and, among influenza A viruses, subtypes (based on the genetic sequences of the hemagglutinin and neuraminidase surface proteins) [21]. Currently, circulating human influenza viruses consist of the A(H1N1) and A(H3N2) influenza subtypes, as well as influenza B viruses [1]. While influenza viruses evolve quickly, significant antigenic change only occurs over a period of one to ten years [22, 23]. However, while the extent of cross-immunity between influenza (sub)types remains unknown, current influenza vaccines are not sufficient to protect people from all (sub)types of influenza virus [21, 24]. Thus, we may expect that separate epidemics within a single year are often caused by different influenza viruses. As a result, appropriate models of influenza transmission in tropical and subtropical locations may need to take multiple types and subtypes into account, further complicating modeling efforts.

Here, we utilize influenza incidence data that have been collected since 1998 in Hong Kong, a densely populated city with a subtropical climate, to explore the impact of climate drivers on influenza epidemics. We formulate seven models, each allowing for differing roles of humidity, temperature, and influenza co-circulation. We expect that, by accounting for 1) increased transmission at both low and high values of AH, 2) decreased transmission at high temperatures (e.g., >30 °C), and 3) co-circulation of several influenza types and subtypes, the model best representing the impact of key climate drivers and influenza co-circulation will be able to best reproduce observed influenza dynamics in Hong Kong. In addition, influenza pandemics could occur “off-season” under more extreme climate conditions (e.g., hot summer days), due to higher population susceptibility to a novel virus. Thus, we include the 2009 pandemic in our model testing and expect the best-performing model to also capture influenza dynamics during the pandemic after accounting for the increased population susceptibility. To model the impact of humidity, here we focus on AH as it is independent of temperature. Specifically, unlike relative humidity measuring the amount of water vapor (i.e. moisture) in the air relative to the total amount of vapor that can exist in the air at its current temperature, AH measures the actual amount of water vapor in the air irrespective of the air’s temperature.

## Methods

### Influenza Data

Data on influenza-like illness (ILI) and laboratory-confirmed influenza from January 1998 through December 2018 were obtained from the Centre for Health Protection of the Hong Kong Special Administrative Region. ILI data were collected by a sentinel surveillance network consisting of 64 public out-patient clinics and roughly 50 private medical practitioners’ clinics throughout Hong Kong, while laboratory testing was performed on specimens from outpatient clinics and public hospitals. Throughout our study period (21 years from January 1998 to December 2018), the same procedure for selecting specimens for viral testing was used; however, from February 10, 2014 onward, viral testing was carried out using molecular testing instead of viral culture. We multiply weekly ILI case counts by the proportion of tests positive for influenza each week and refer to the resulting, more specific measure as ILI+. Finally, data were converted to rates per 100,000 population.

### Climate Data

Hong Kong has a humid subtropical climate, with hot, humid, and rainy summers, and mild winters [25]. Daily mean temperature and relative humidity were taken at the Hong Kong Observatory [26]. Using these data, we used the Clausius-Clapeyron relation [27] to calculate daily mean specific humidity, a measure of AH (see Supplementary Materials).

### Model Hypotheses

In this work, we formulated seven models to test several hypotheses concerning the impact of humidity and temperature on influenza transmission in Hong Kong, and how co-circulation influenza (sub)types affect this process. While specific methodology associated with each model is detailed below, we describe the hypotheses considered here:

- Null hypothesis 1 (or Null1, Constant basic reproductive number *R*_*0*_): Climate conditions have no effect on influenza transmission. To represent this, we held *R*_*0*_, the epidemic parameter representing the average number of new cases arising from a primary case in a fully susceptible population, constant.
- Null hypothesis 2 (or Null2, Temperate Forcing): AH affects influenza transmission in the subtropics and tropics in the same manner as in temperate regions; that is, *R*_*0*_ increases monotonically with decreasing AH. Here, *R*_*0*_ was modeled according to the equation first presented in [11]. This model has previously been shown to perform well when modeling influenza transmission patterns in temperate regions [12].
- AH model: The effect of AH is bimodal, with both low and high AH conditions favoring influenza transmission. Temperature is assumed to have no additional effect on transmission. *R*_*0*_ was modeled according to Eqn S6, where both low and high values of AH lead to higher values of *R*_*0*_.
- AH/T model: Absolute humidity has a bimodal effect on *R*_*0*_, as in the AH model, but this effect is moderated by temperature as shown in Equation 3. Briefly, low temperatures promote influenza transmission, and temperatures above a certain threshold limit transmission.
- AH/T/Strain: AH and temperature impact *R*_*0*_ as in the AH/T model. Additionally, two “strains” of influenza co-circulate in the population, as in [12]. Here we ignore cross-immunity between strains; in other words, infection with one strain has no effect on the potential for later infection with the other strain.
- AH/T/Short: Climate forcing is included as in the AH/T model. Additionally, we restrict the duration of immunity in the model to be about 1 year (i.e., 0.5 – 1.5 years), in order to implicitly take co-circulation into account (i.e., for an individual, multiple infections by different influenza strains could occur within a short time span).
- AH/T/Vary: Climate forcing is included as in the AH/T model. Additionally, a small proportion (<50%) of the population experiences a significantly truncated duration of immunity (<1 year) after infection. In other words, duration of immunity to influenza is heterogeneous among the population.

### SIRS Model

We modeled influenza transmission in Hong Kong using a compartmental susceptible-infected-recovered-susceptible (SIRS) model with demography. While we used two distinct forms of the model for this project described below, the basic model takes the form:

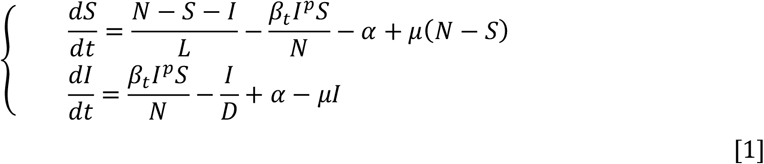

where *N* is the size of the model population (here set to 100,000); *β*_*t*_ represents the rate of transmission on day *t*, depending on climatic functions as described below; *D* is the mean infectious period; *L* is the average duration of immunity; and *α* represents random seeding from outside the model population (here we arbitrarily set it to 0.1, i.e. 1 per 10 days for all models). *µ* is the rate of natural birth and death (i.e., to maintain a constant population size, we assume equal birth and death rates). We set *µ* to be 0.00918 divided by 365, or the average daily birth rate per person in Hong Kong during 1998 – 2017 [28]. The parameter *p* is an exponent to introduce nonlinearity into the infection process (i.e. imperfect population mixing). The inclusion of this parameter has been shown to be helpful in modeling complicated epidemics using very simple models [29], as is the case in this work. Finally, because the 2009 pandemic was caused by a novel strain of influenza with little prior population immunity, we reset the number of susceptibles in our population in early August of 2009 to be between 60% and 80% of the total population [30-32] (vs. 40-80% for model initiation in January 1998; see Table 1). All models were run stochastically, as described in the Supplementary Materials and in [12].

**Table 1.** Descriptions and estimates of the parameter ranges. The estimated parameter ranges are the 95% highest density intervals, obtained after two rounds of parameter selection from the initial ranges (shown in parentheses).

| *Parameter and sources of initial ranges* | Parameter Description | Null1 | Null2 | AH | AH/T | AH/T/Strain | AH/T/Short | AH/T/Vary |
| --- | --- | --- | --- | --- | --- | --- | --- | --- |
| $R_0$<br>[12, 34, 36-38] | The basic reproductive number (i.e., the average number of cases caused by a primary case in a fully susceptible population). | 2.06-2.11<br>(1.0-3.0) | NA | NA | NA | NA | NA | NA |
| $R_{0mx}$<br>[12, 34, 36-38] | The theoretical value of $R_0$ at $q = q_{min}$ and $q = q_{max}$, when $T = T_c$ | NA | 2.52-2.69<br>(1.5-3.0) | 1.95-2.50<br>(1.5-3.0) | 2.34-2.93<br>(1.5-3.0) | 2.38-2.88<br>(1.5-3.0) | 2.13-2.67<br>(1.5-3.0) | 2.27-2.66<br>(1.5-3.0) |
| $R_{0diff}$ | The difference between $R_{0max}$, and $R_0$ at $q = q_{mid}$. | NA | 1.10-1.20<br>(0.6-1.2) | 0.60-1.18<br>(0.6-1.2) | 0.86-1.18<br>(0.6-1.2) | 0.70-1.17<br>(0.6-1.2) | 0.65-1.09<br>(0.6-1.2) | 0.72-1.12<br>(0.6-1.2) |
| $q_{min}$ (g/kg) | The absolute humidity value at which $R_0 = R_{0max}$ when $T = T_c$; the minimum value of absolute humidity permitted. | NA | NA | 7.7-8.0<br>(2.0-8.0) | 2.2-4.0<br>(2.0-8.0) | 2.0-3.6<br>(2.0-8.0) | 2.1-3.4<br>(2.0-8.0) | 2.1-3.7<br>(2.0-8.0) |
| $q_{max}$ (g/kg) | The absolute humidity value at which $R_0 = R_{0max}$ when $T = T_c$; the maximum value of absolute humidity permitted. | NA | NA | 22.6-23.0<br>(16.0-23.0) | 17.0-20.0<br>(16.0-23.0) | 17.0-19.0<br>(16.0-23.0) | 18.0-19.0<br>(16.0-23.0) | 17.0-19.0<br>(16.0-23.0) |
| $q_{mid}$ (g/kg) | The absolute humidity value at which $R_0 = R_{0max} - R_{0diff}$. | NA | NA | 12.6-13.0<br>(10.0-13.0) | 10.2-11.3<br>(10.0-13.0) | 10.2-12.5<br>(10.0-13.0) | 10-12.2<br>(10.0-13.0) | 10.2-12.6<br>(10.0-13.0) |
| $T_c$ (°C)<br>[33, 35] | The cutoff temperature above which temperature negatively impacts $R_0$. | NA | NA | NA | 20.24-24.0<br>(20-25) | 20.04-24.46<br>(20-25) | 20.16-24.47<br>(20-25) | 20.52-23.87<br>(20-25) |
| $T_{diff}$ (°C) | A parameter that, when subtracted from $T_c$, yields the minimum temperature permitted as a model input. | NA | NA | NA | 0.40-5.09<br>(0-15) | 1.28-7.05 (0-15) | 1.34-5.31 (0-15) | 0.77-4.46 (0-15) |
| $T_{exp}$ | An exponent determining the strength of the impact of temperature on $R_0$. | NA | NA | NA | 0.95-1.54<br>(0.5-2.0) | 0.67-1.49<br>(0.5-2.0) | 0.78-1.33<br>(0.5-2.0) | 0.78-1.34<br>(0.5-2.0) |
| $D$ (days)<br>[12, 30, 31, 38] | The duration of influenza infection. | 3.35-3.71<br>(2-5) | 4.68-4.98<br>(2-5) | 2.56-4.99<br>(2-5) | 4.05-4.99<br>(2-5) | 3.50-4.70<br>(2-5) | 3.87-4.98<br>(2-5) | 3.67-4.90<br>(2-5) |
| $L$ (days)<br>[12, 22, 23, 36, 38] | The duration of influenza immunity; in the AH/T/Vary model, the duration of immunity among those who do not lose immunity at an accelerated rate. | 369-391<br>(365-3650) | 421-607<br>(365-3650) | 1298-2860<br>(365-3650) | 376-489<br>(365-3650) | 536-838<br>(365-3650) | 310-451<br>(183-548) | 374-567<br>(365-3650) |
| $L_s$ (days) | The duration of influenza immunity among those with accelerated immunity loss in the AH/T/Vary model. | NA | NA | NA | NA | NA | NA | 82-336<br>(30-365) |
| $\rho$ | The proportion of the population with accelerated immunity loss in the AH/T/Vary model. | NA | NA | NA | NA | NA | NA | 0.0006-0.29<br>(0-0.5) |
| $S_0$ | The number of people susceptible to influenza at the beginning of the model run. | 48.78%<br>-<br>76.16% | 78.77%-79.98%<br>(40%-80%) | 71.58%<br>-<br>79.71% | 59.27%<br>-<br>76.79% | 57.03%<br>-<br>69.86% | 67.03%<br>-<br>77.61% | 66.02%-79.36<br>(40%-80%) |
|  |  | (40%-80%) |  | (40%-80%) | (40%-80%) | (40%-80%) | (40%-80%) |  |
| $I_0$ | The number of people infected at the beginning of the model run. | 520-1489<br>(500-1500) | 534-851<br>(500-1500) | 611-1161<br>(500-1500) | 515-1052<br>(500-1500) | 512-1105<br>(500-1500) | 515-900<br>(500-1500) | 519-1256<br>(500-1500) |
| $p$ | An exponent to allow for imperfect mixing. Homogenous mixing occurs when $p = 1.0$. | 0.97 | 0.97 | 0.97 | 0.97 | 0.97 | 0.97 | 0.97 |

### SIRS Model Variations

In this work, we considered three models to account for cocirculation and heterogenous immunity period:

1. **AH/T/Strain:** To account for co-circulating influenza viruses, we ran two SIRS simulations (Eqn 1) in parallel, as done in Shaman et al. [12]. While there are three co-circulating influenza (sub)types, we combined A(H1N1) and B for simplicity, given the similar timing of the circulation of these two viruses (Fig S1). Here, we applied random seeding at each week only to the “strain” that had the greater number of positive tests that week. If both “strains” were equally common, seeding was applied to a single strain at random. We then combined the output of the two simulations and compared the resulting estimates to the overall ILI+ data. Note that per this simple model, multiple influenza epidemics due to different influenza viruses can occur at the same time, as well as co-infection of multiple influenza viruses.
2. **AH/T/Short:** This model used Equation 1 but restricted *L*, the duration of immunity, to be between around one year to account for multiple infections within a year due to multiple circulating strains.
3. **AH/T/Vary:** In this model, a small proportion of the population loses immunity to influenza at an accelerated rate. This was modeled by replacing the term 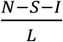 in Equation 1 with:

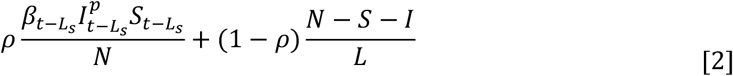

where *ρ* is the proportion of the model population that loses immunity after a short period *L*_*s*_ and 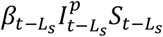 represents the number of new infections *L*_*s*_ days ago, who would lose their immunity on day *t*. We note that, since longer-term immunity is lost at an exponential rate, while short term immunity simply removes a set number of individuals from the recovered compartment at each time step, there is a possibility of double-counting. However, given the difference in time scales (less than one year vs. several years), we do not anticipate that this approximation will lead to severe problems.

### Climate Forcing Models

Based on past work [12, 14, 16, 20], as well as the patterns observed in the influenza and climate data described above and shown in Fig 1, we modeled the impact of AH using a parabola, where transmissibility is highest at very low and very high levels of AH. In all AH/T models, this relationship was modified by temperature such that, when temperature is above some cutoff value, transmissibility is reduced. Specifically, for models AH/T, AH/T/Strain, AH/T/Short, and AH/T/Vary, the impact of AH and temperature on transmissibility was modeled as:

**Fig 1.**
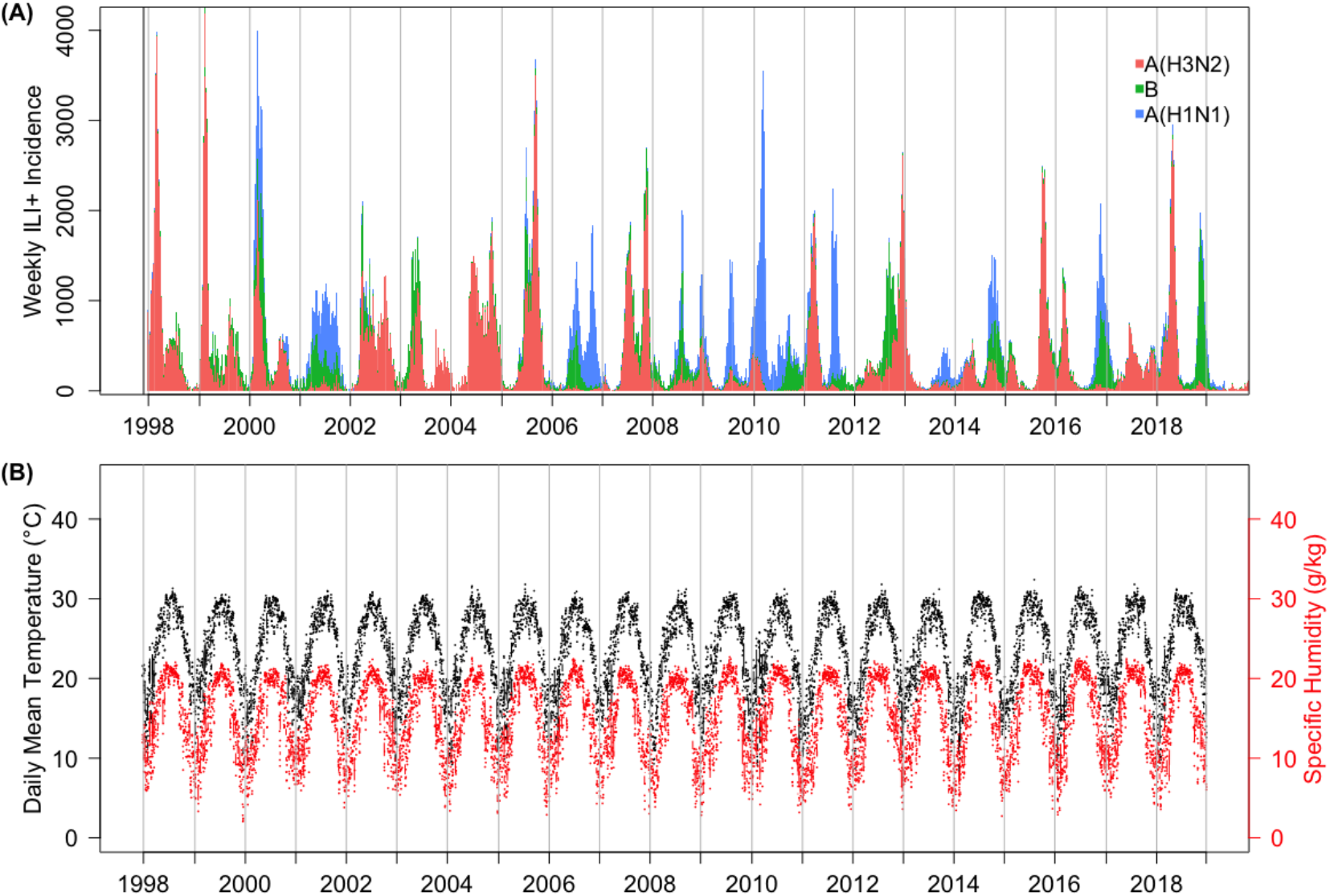
Influenza epidemics observed in Hong Kong during 1998-2018 and the corresponding mean daily temperature and AH. Upper panel: stacked barplot of the weekly ILI+ incidence time series. Segments of the bar represent different virus (sub)types circulating during the week. Lower panel: daily mean temperature and specific humidity (a measure of AH) observed during 1998-2018.

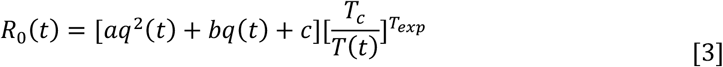

where *q(t)* is specific humidity (i.e. a measure of AH) at time *t*, and *T(t)* is temperature at time *t*. The parameter *β*_*t*_ in the SIRS model is defined as *R*_*0*_*(t)* divided by *D;* thus, AH and temperature act through *β* in Equations 1 and 2 to influence transmission patterns. When *T* is below *T*_*c*_, lower temperatures are able to further increase *R*_*0*_, whereas temperatures above *T*_*c*_ inhibit influenza transmission. However, the favorable impact of low temperature may level off at relatively low temperatures. Thus, we truncated this monotonic relationship at a minimum temperature of *T*_*c*_ *–T*_*diff*_, beyond which, the effect levels off. The strength of this relationship is further determined by the exponent *T*_*exp*_. For the AH Only model, AH was also modeled as a parabola (i.e. the terms within the first set of squared brackets), but with no impact of temperature (Eqn. S6).

To link the coefficients *a, b*, and *c* to the AH *q* and *R*_*0*_, we reparametrize them by solving the parabola with the nadir at (*q*_*mid*_, *R*_*0min*_) and maximum at both (*q*_*min*_, *R*_*0max*_) and (*q*_*max*_, *R*_*0max*_) (see Supplemental Materials).

### Model optimization (parameter tuning)

To assess the validity of the models, we first split the influenza data into a training set (Jan 1998 – Dec 2012; 15 years) and a testing set (Jan 2013 – Dec 2018; 6 years). That is, data from 1998 to 2012 were used to train the model and optimize parameters, the remaining data from 2013 to 2018 were holdout for testing.

We optimized each model by testing a wide range of parameter values (Table 1) based on the literature [12, 22, 23, 33-39], and tuning parameter range against the influenza data observed during Jan 1998 – Dec 2012 (i.e. the training period). Specifically, for each model, we draw 1 million parameter combinations from ranges listed in Table 1 using Latin hypercube sampling [12]. Using each parameter combination, we ran each model stochastically from Jan 1998 to Dec 2012 with a daily time step and aggregated the simulated daily ILI+ to weekly intervals for model assessment. To account for model stochasticity, we repeated the simulation 500 times for each parameter set. We then calculated, for each model run, the corresponding root mean square error (RMSE) and correlation against the full training dataset and weekly averaged dataset (i.e., for each of the 52 weeks of the year, averaged the ILI+ over 15 training years), separately; We refer to these metrics as *full*.*RMSE, avg*.*RMSE, full*.*Correlation* and *avg*.*Correlation*, respectively, hereafter. We further averaged across the 500 model runs for each parameter combination to obtain a single set of metrics for the corresponding parameter combination. To combine the metrics and simplify the process of parameter selection, we averaged the two RMSE metrics (RMSES) and the two correlation metrics (CORR), separately, as:

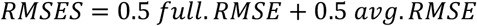

and:

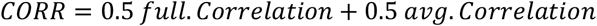

Based on these two final metrics, we selected the top 1000 parameter combinations out of the one million for each model: the 1000 with the highest *CORR* among those with RMSES lower than the 0.5 percentile. We then computed the 95% highest density interval (HDI) for each parameter using those top 1000 to generate new parameter ranges for subsequent round of optimization. That is, we updated the range of a parameter with the HDI, if the upper (or lower) bound of HDI is 10% smaller (or larger) than the corresponding bound of the original range. This tuning process was repeated until the parameter range no longer shrinks substantially (i.e. by 10%). In this study, it took two rounds of such parameter selection; at each round, we checked to ensure the model fit was improving using the new parameter ranges.

After obtaining the final parameter ranges, we drew 10,000 random parameter combinations from these final ranges and ran the models from Jan 1998 – Dec 2018 using each parameter combination. In this last batch, we obtained the top 1000 parameter combinations as described above for further assessment and model comparison.

### Model assessment

We evaluated performance of the seven models based on the four metrics described above (i.e., *full*.*RMSE, avg*.*RMSE, full*.*Correlation, avg*.*Correlation*) during both the training and testing period. To quantify the differences in performance among models, we applied the Kruskal-Wallis test to test if the mean rank of different models were similar for each metric. Since significant differences were found between models, we further performed a pairwise comparison using the Nemenyi test with an adjusted p-value of 0.007 (i.e., 0.05 /7). We summarized the model ranking in Table 2 based on model performances including the pandemic period. In addition, as a sensitivity analysis, we ranked the models based on their performances excluding the 2009 pandemic in Table S1.

**Table 2.** The model performance ranks for each comparison metric using training and testing data. The rankings are determined by the model’s absolute mean rank differences with the best-ranked model. Different rankings between models indicate significantly different absolute mean ranks (i.e., p-value<0.007). The mean value of the corresponding metric is shown in the parenthesis.

|  | Models | Null1 | Null2 | AH | AH/T | AH/T/<br>Strain | AH/T/<br>Short | AH/T/<br>Vary |
| --- | --- | --- | --- | --- | --- | --- | --- | --- |
| Train | full.RMSE | 6<br>(772) | 4<br>(675) | 7<br>(804) | 1<br>(611) | 6<br>(720) | 1<br>(604) | 1<br>(611) |
|  | avg.RMSE | 6<br>(422) | 5<br>(349) | 7<br>(541) | 1<br>(243) | 4<br>(310) | 1<br>(240) | 1<br>(249) |
|  | full.Correlation | 7<br>(0.12) | 5<br>(0.43) | 6<br>(0.32) | 2<br>(0.53) | 4<br>(0.50) | 1<br>(0.55) | 2<br>(0.52) |
|  | avg.Correlation | 7<br>(-0.16) | 5<br>(0.73) | 6<br>(0.62) | 3<br>(0.87) | 1<br>(0.88) | 1<br>(0.89) | 3<br>(0.87) |
|  | Average rank | 6.5 | 4.75 | 6.5 | 1.75 | 3.75 | 1 | 1.75 |
| Test | full.RMSE | 6<br>(620) | 3<br>(478) | 5<br>(613) | 2<br>(479) | 7<br>(676) | 3<br>(492) | 1<br>(470) |
|  | avg.RMSE | 6<br>(434) | 2<br>(237) | 4<br>(422) | 2<br>(249) | 6<br>(476) | 4<br>(274) | 1<br>(228) |
|  | full.Correlation | 7<br>(-0.0005) | 5<br>(0.49) | 6<br>(0.10) | 3<br>(0.54) | 1<br>(0.55) | 1<br>(0.54) | 4<br>(0.52) |
|  | avg.Correlation | 7<br>(-0.0007) | 5<br>(0.76) | 6<br>(0.11) | 1<br>(0.78) | 1<br>(0.78) | 1<br>(0.79) | 4<br>(0.78) |
|  | Average rank | 6.5 | 3.75 | 5.25 | 2 | 3.75 | 2.25 | 2.5 |

## Results

### General influenza transmission dynamics in Hong Kong

Influenza activity is highly diverse in Hong Kong. Unlike the common wintertime epidemics in temperate regions, during our study period (Jan 1998– Dec 2018), Hong Kong experienced two epidemics per year in most years (17 out of 21; the four exceptions were: Year 2001, 2011, 2012 and 2018; Fig 1A). For years with two epidemics, one epidemic typically occurred during the winter, and the other, usually smaller in magnitude, occurred during the summer (Fig 1A). Usually, there was a single influenza (sub)type predominating during each epidemic, although co-circulation of multiple strains with comparable magnitudes could occur (Fig 1A). While any (sub)type can dominate winter epidemics, summer epidemics were almost always caused by A(H3N2) (15 out of 17 years with summer epidemics; Fig 1A).

### Climate conditions in Hong Kong overall and during influenza epidemics

Both mean daily temperature and AH in Hong Kong displayed strong seasonality, where temperature and AH were highest in summer and lowest in winter (Fig 1B). In the hottest days, the mean temperature in Hong Kong can exceed 30°C (around six days per year), but most of the time, mean temperature was between 15°C to 30°C. Hong Kong was also very humid. Compared to the relatively drier and cooler climates in temperate regions, the humid climate in Hong Kong may provide favorable conditions for influenza transmission during the summer despite the high temperatures. In addition, temperature and AH were highly correlated (Pearson’s *ρ* = 0.94, *p* < 0.001). Such a high correlation could confound the potential effect on influenza activity due to either climate variable and suggests that simple linear models will not be able to separate such effects.

### Models incorporating the impact of humidity and temperature best replicate the influenza dynamic in Hong Kong

In this work, we designed seven models and performed two rounds of parameter selection to obtain the optimized HDI for each parameter in every model, and eventually compared the models using the selected parameter combinations. Among the seven models constructed, the Null1 model served as a “control” model since it had a constant *R*_*0*_ that is independent of any climate factors. In other words, if a model failed to outperform the Null1 model, then the proposed relationship between climate factors and the influenza transmission dynamics would not be supported. In addition, we note, again, that we included the 2009 influenza pandemic in our main analysis, because the off-season occurrence of pandemic influenza provided a unique opportunity to test the climate forcing models under more extreme climate conditions. Table 2 shows the model ranking including the pandemic and Table S1 excluding the pandemic. Results are consistent across the two analyses.

Overall, all models incorporating seasonality outperformed the Null1 model, for which *R*_*0*_ is set to a constant (mean testing rank: 6.5 out of seven models; Table 2 and Fig 2), suggesting the seasonal variation of influenza activity. In addition, while the AH model (i.e. without including temperature as a variable) outperformed the Null1 model, it was consistently inferior when compared to other models (mean testing rank: 5.5 out of 7 models; Table 2, Fig 2). The model failed to capture much of the dynamics observed in Hong Kong, especially in colder months (Fig S2C). This suggests that, in addition to AH, temperate can modulate influenza transmission in Hong Kong.

**Fig 2.**
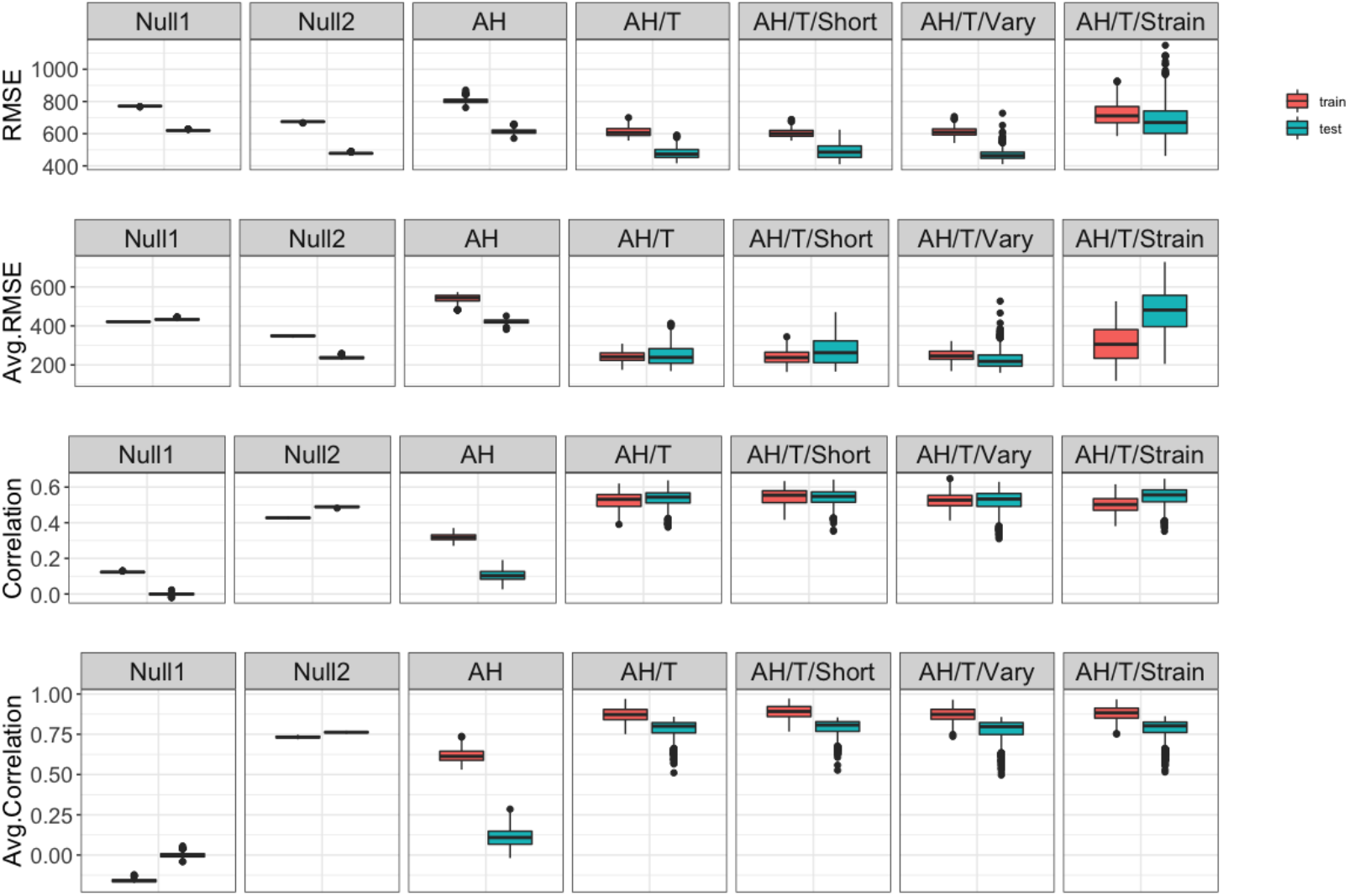
Model performance. Boxes and whiskers show the median (thick horizontal lines), interquartile range and 95% CI of RMSE (1^st^ row), average RMSE (2^nd^ row), correlation (3^rd^ row) and average correlation (4^th^ row) of the top 1000 parameter combinations for each model, during the training (red) and testing (green) period, separately.

To test if the monotonic increase in transmission with decreasing AH as observed in temperate regions also applies to the subtropics, we also tested the model by Shaman et al. (i.e. the Null2 model here). The Null2 model was able to better recreate the observed transmission dynamics in Hong Kong than the Null1 and AH model. However, its performance during the training period ranked lower than models additionally incorporating increasing transmission under high AH conditions (i.e. a bimodal relation) (Table 2).

Models incorporating the impact of both AH and temperature improved the model performance significantly (Fig 2). The AH/T models (AH/T, AH/T/Short, AH/T/Vary, and AH/T/Strain) had the best performance among all. Among these four models, the AH/T and AH/T/Short models assumed the same model construct but with different initial ranges for the immunity duration (*L*). After rounds of parameter selection, the HDI of *L* eventually converged for the two models (Table 1), as did other parameters. Therefore, we considered these two models to be the same. Indeed, either model ranked among the top two during the training period. However, the AH/T model appeared to be more stable and remained as a top performing model during the testing period (Table 2).

Two models (i.e., AH/T/Vary and AH/T/Strain) additionally accounted for long/short-term immunity and/or co-circulation of different influenza viruses. Both models performed comparably to the AH/T model but not better (Fig 3). In addition, for the common parameters, the HDIs of the three models converged to similar ranges (Table 1).

**Fig 3.**
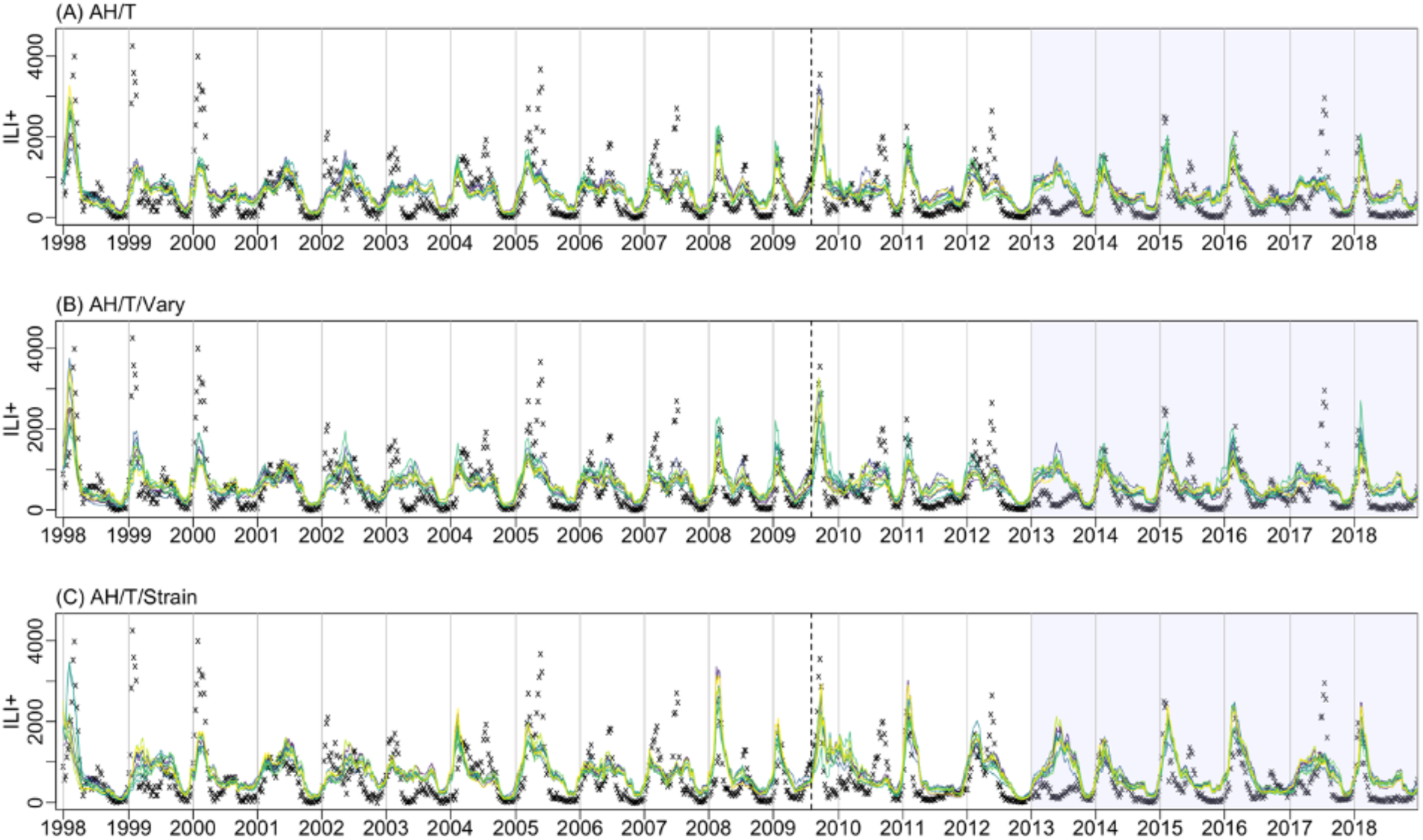
Top10 model fits for three climate forcing models: AH/T (A), AH/T/Vary (B), and AH/T/Strain (C). Black crosses show observed ILI+; the colored lines run through the crosses are the top10 model estimates. The vertical dash line indicates a pandemic (2009). The shaded region represents testing years (2013-2018), while the rest are the training years.

The AH/T/Vary model, although formulated differently, had similar HDI and performances as the AH/T model. The AH/T/Strain model was the only model that specifically accounted for co-circulation of different influenza viruses. Although the model was able to simulate ILI+ with high correlation with the observed data and recreate large epidemics, it did not fit smaller epidemics as well and thus had a high RMSE (Fig 3).

Taken together, the AH/T model had the most consistent and best performance over the entire study period. It is also the most parsimonious model among all top performing ones.

### Impact of humidity and temperature on influenza transmission, as estimated by the top models

To account for the impact of humidity on influenza transmission, we included three parameters in the models to describe a bimodal relationship. That is, below a threshold AH (*q*_*mid*_), transmission would increase as AH decreases and level off when AH is ≤ *q*_*min*_, whereas above that threshold, transmission would increase as AH increases and level off when AH is ≥ *q*_*max*_. Tamerius et al 2013 [14] defined a similar bimodal relationship and found a threshold of 11-12 g/kg for regions with single-peak vs bimodal epidemics. In addition, they found that most influenza peak activities occurred in “cold-dry” conditions (i.e., AH < 8 g/kg) or “humid-rainy” conditions (AH > 14 g/kg). Consistent with Tamerius et al., we estimated the threshold *q*_*mid*_ to be 10 – 12.6 g/kg, where *R*_*0*_ troughs (Fig 4). In addition, we further quantified how influenza transmission (i.e. as indicated by *R*_*0*_ estimates) changes with AH in the two regimes (Eqn 3). We estimated that when AH is below 10-12.6 g/kg, *R*_*0*_ would increase with decreasing AH quadratically up to a minimum of 2 – 4 g/kg; when AH is above 10-12.6 g/kg, *R*_*0*_ would increase with increasing AH quadratically up to a maximum of 17-20 g/kg (Table 1).

**Fig 4.**
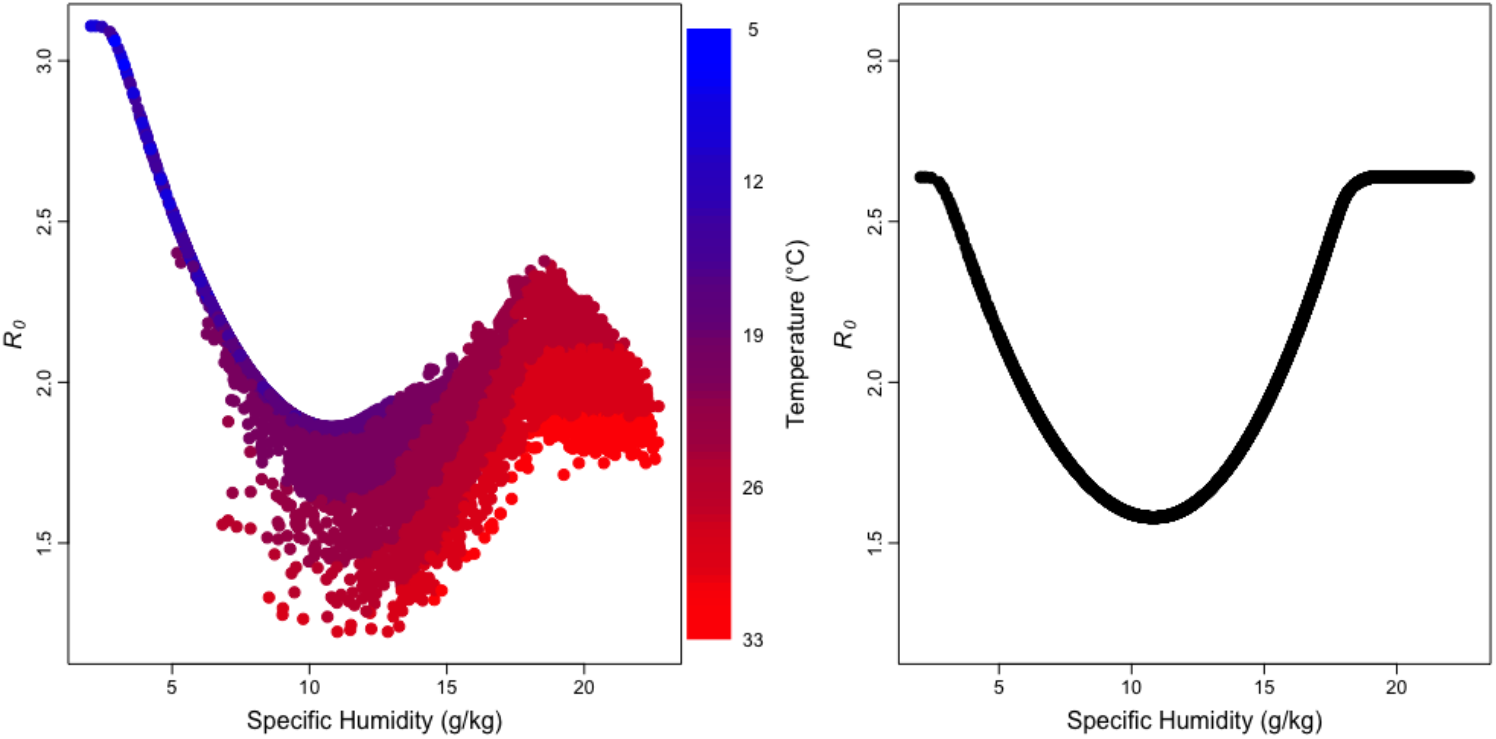
Estimated relationship between influenza transmission with AH and temperature. We use the basic reproductive number (*R*_*0*_) to represent the level of influenza transmission. Each point shows the estimated *R*_*0*_ at different specific humidity, a measure of AH, (and temperature if included) calculated per the AH/T model (left) or the AH model (right) using the top 10 parameter combinations for the corresponding model. For the AH/T model (left panel), the color of the point shows the concurrent temperature included in the model to moderate the relationship between *R*_*0*_ and specific humidity.

In addition to AH, we included three parameters to model the impact of temperature on influenza transmission. Foremost, as described in Eqn 3, the impact of temperature is modeled as a multiplicative adjustment to the impact of AH. Below a threshold temperature *T*_*c*_, decreasing temperature increases transmission up to a minimum of *T*_*c*_ *–T*_*diff*_; above *T*_*c*_, increasing temperature reduces transmission. Deyle et al. [20] suggested temperature around 24°C (75°F) as a threshold dividing the negative and positive effect of AH on transmission. Similarly, we estimated *T*_*c*_ to have an HDI between 20-24°C (Table 1). As shown in Fig 4, when temperature is below *T*_*c*_, low temperature further facilitates transmission, in addition to the favorable transmission conditions due to concurrent, low AH in cold months. However, when temperature exceeds *T*_*c*_, high temperature suppresses transmission and lowers the overall *R*_*0*_ despite the favorable transmission conditions due to concurrent, very high AH in the summer. In addition, we estimated *T*_*diff*_ to have an HDI of 0.4-5.3°C, which implies temperature below 16-23°C does not afford additional increases in transmission. Similar transmission behavior was also found by Brown et al. [33] with avian influenza viruses, whose infectiveness stabilized after the temperature dropped below 17°C. The estimated value of *T*_*exp*_, the exponent of the *T*_*c*_/*T* ratio (Eqn 3), had an HDI of 0.95-1.54, suggesting the moderation of temperature is slightly super-linear.

We also estimated the duration of immunity (*L*) with our models to better understand transmission dynamics. The immunity period *L* directly affects the frequency of influenza epidemics – i.e., shorter *L* would lead to more frequent epidemics. Here we estimated *L* to be around 1-1.3 years for the AH/T model, and 1.5-2.3 year for AH/T/Strain. For the AH/T/Vary model, considering population heterogeneity, we included another parameter *L*_s_ to represent short-term immunity (<1 year). We found *L*_*s*_ was around 3-11 months and *L* around 1-1.6 years; combing these two parameters (i.e., *L, Ls*), the average is very similar to the *L* estimated by the AH/T model. Overall, these *L* estimates are consistent with the frequent influenza epidemics observed in Hong Kong; however, we note that these estimates are on the lower end of the reported 1-10 year range [22].

### Validate model findings using laboratory results

The reproductive number represents the epidemic potential of an infection. To examine if the identified relationship with AH and temperature is consistent with data from laboratory studies, we further compared the basic reproductive number *R*_*0*_ and effective reproductive number *R*_*e*_ calculated by our model (Eq 3) with the influenza virus survival rate in aerosols [35] as well as the influenza transmission rate observed in guinea pigs [39, 40]. At the population level, epidemic can occur when *R*_*e*_ is above 1 and will subside when *R*_*e*_ drops below 1. Indeed, no transmission occurred in the guinea pig studies [40] under conditions where the estimated *R*_*e*_ per our model (Eqn 3) was below 1; in contrast, transmission occurred when estimated *R*_*e*_ was above 1, ranging 1.21-2.25 (corresponding *R*_*0*_ range: 1.68-3.11; Fig. 5). In addition, fitting a linear regression of estimated *R*_*0*_ (or *R*_*e*_) against the log survival rate and log transmission probability, we found that estimated *R*_*0*_ (or *R*_*e*_) positively correlated with both laboratory-observed survival rate and transmission rate of the influenza virus (Fig 5). In particular, the correlation between *R*_*0*_ (or *R*_*e*_) estimated by our AH and temperature model (Eqn 3) and the observed transmission rate was 0.82 (*r*^2^ = 0.67; Fig 5), suggesting it was able to explain around 70% of the variances of the transmission rate. These findings indicate that our model is able to represent both the impact of AH and temperature on influenza epidemic patterns at the population level as well as influenza transmission and survival observed in laboratory settings.

**Fig 5.**
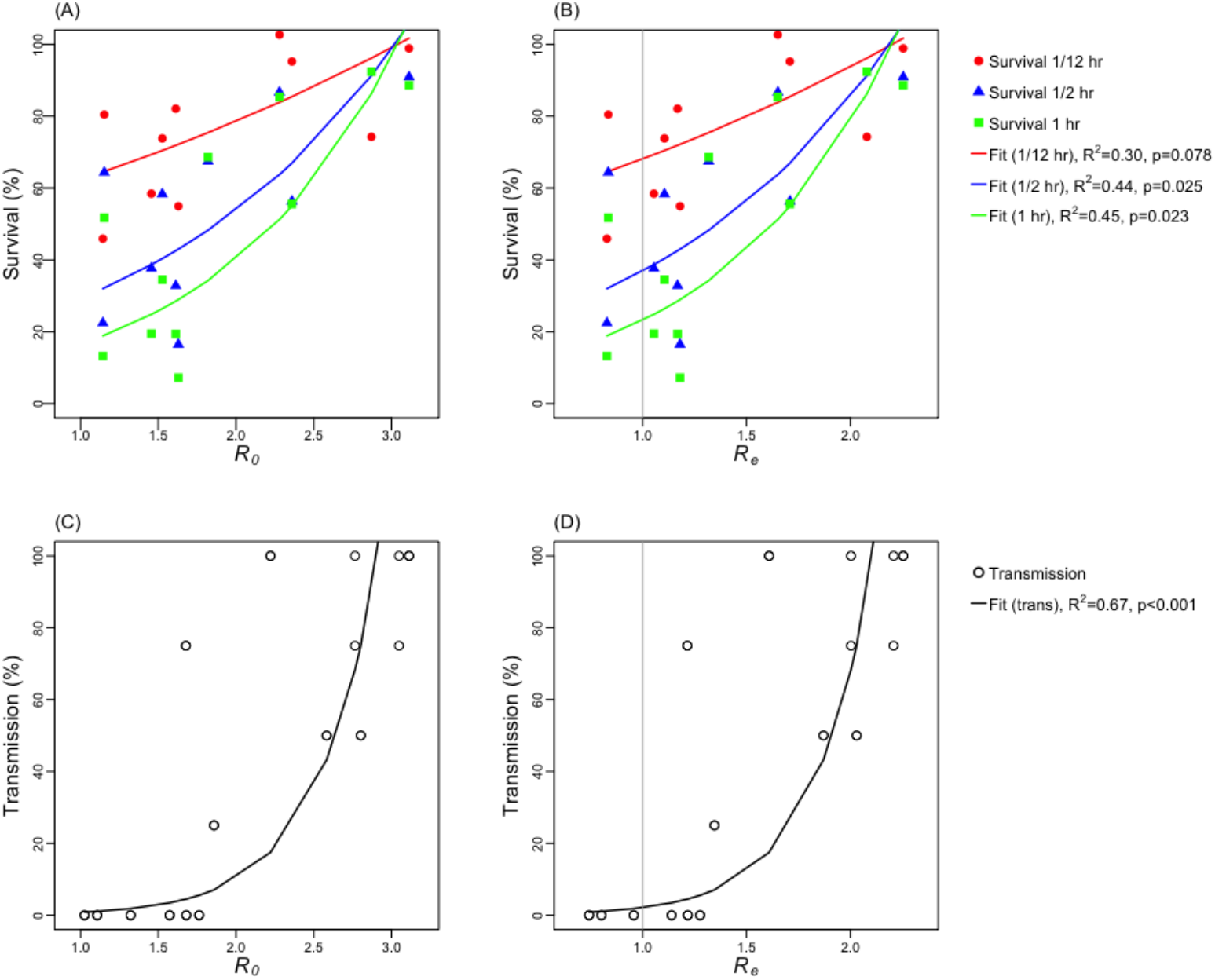
Comparison of the reproductive numbers estimated by the AH/T model with laboratory observed virus survival rate and transmission rate in guinea pigs. Left panel plots the viral survival rate (A) and transmission rate (C) against *R*_*0*_ calculated using Eqn 3 and best-fit parameters for the AH/T model. Right panel plots the viral survival rate (B) and transmission rate (D) against *R*_*e*_, where *R*_*e*_ is calculated as *R*_*0*_ multiplied by the estimated mean population susceptibility during the study period. The grey vertical line indicates where *R*_*e*_ =1. The viral survival data came from Harper 1961 [35] and transmission rate data came from Lowen et al. 2007 and 2008 [39, 40].

## Discussion

Despite influenza’s profound impact on public health, little is known about how the virus transmits from person to person and what environmental and climate conditions make this process more likely. To date, there have been only a few studies on the influenza burden in tropical and subtropical regions. Of those studies, a handful analyzed the effect of climate drivers on influenza incidence using time-series or logistic regression models. Although those statistical models were able to identify significant climate covariates (including AH, temperature, and precipitation) and estimate the effect size, they did not provide much information to the underlying mechanism of how those climate covariates affect transmission. In contrast, infectious disease models (e.g., the SIRS model) provide a means to model and test the relationship between climate factors and influenza transmission observed in laboratories. In this work, by building seven SIRS models under different hypotheses, we explored how temperature, AH, and influenza co-circulation affect the transmission of influenza. We showed that models that included both AH and temperature as covariates consistently outperformed those that did not in recreating the observed influenza epidemic patterns in Hong Kong. These results support that climate variables play a critical role in influenza transmission, where temperature is particularly influential in moderating the impact of AH. Model results also indicate that the effect of climate drivers in tropical and sub-tropical regions is different from those observed in the temperate regions, and models built for temperate areas will not be sufficient to reproduce the transmission patterns in the tropics or the subtropics.

Previous studies had suggested a bimodal effect of AH on influenza transmission [20]. However, such an effect has not been quantified nor incorporated in influenza transmission models as done for temperate regions [12]. Here, we have developed a model that can effectively represent this U-shaped relationship with AH, moderated further by temperature. When incorporated into an SIRS model, the combined model was able to recreate the diverse influenza epidemic dynamics observed in Hong Kong over a 21-year period. As described earlier, the breaking point between the “cold-dry” and “hot-humid” environment is an AH of approximately 10-12.6 g/kg, which is consistent with Tamerius et al 2013 [14]. This finding helps to explain the biannual epidemics observed in Hong Kong. In the winter when weather is “cold-dry,” the influenza transmission rate increases as the environment gets drier. On the other hand, summer in Hong Kong is hot and humid – the very high AH also promotes influenza transmission, despite high temperatures.

The combined relationship of influenza transmission with both AH and temperature we delineated in Fig 4 also consistently explains observations in temperate regions where influenza surges predominantly in winter and increases monotonically with decreasing AH. During summers in temperate areas, absolute AH is typically lower than 15 g/kg (vs. as high as 23g/kg in subtropics/tropics) [12], and temperature is relatively high. Under such environmental conditions, *R*_*0*_ would stay in the trough during the summer (Fig 4), and only increases in the winter when AH becomes low (Fig 4). As such, it appears that influenza transmission rate increases monotonically with decreasing AH, leading to a single epidemic each year in the winter in temperate regions.

When there are multiple strains co-circulating, individuals who recover from infection gain specific immunity against the infecting strain and may remain susceptible to other influenza strains in co-circulation either during the same or subsequent epidemics. In this study, we combined data from all three co-circulating influenza viruses and, as such, our estimated duration of immunity, without distinguishing specific strains, was relatively short (∼1–1.5 years). In comparison, the AH/T/Strain model which modeled H3N2 and H1N1/B epidemics separately, estimated a longer immunity period (1.5-2.5 years). Nevertheless, either estimate is quite low compared to the duration an influenza clade in circulation (1-10 years). Therefore, we note that the short immunity duration estimated here is more to capture the frequent influenza epidemics in Hong Kong. For locations with less frequent influenza epidemics, re-estimating the immunity duration per local epidemic data is warranted.

We recognize several limitations of our study. First, although our models fit the observations in Hong Kong well and provide support to the role of AH and temperature on influenza transmission, it is important to note that further laboratory and epidemiologic works are needed to establish a causal relationship. Second, while we accounted for multi-strain co-circulation to some extent (e.g., the AH/T/Short and the AH/T/Strain model), our models are highly simplified and do not distinguish the different epidemiological characteristics for H3N2, H1N1, and B; as such, our estimate of the immunity period may not reflect the true value. Third, for model simplicity, we did not model the interactions between (sub)types, which can modify the association with climate variables [41]. Future research is necessary to explore the impact of co-circulation, as a reasonable inclusion of multiple influenza (sub)types may be more appropriate than combining all strains. Fourth, as a first step, here we combined all influenza viruses without separating influenza by subtype or type. Previous studies have reported differential responses of different influenza strains to climate variables [41]. Future work could also model the impact of climate variables for each influenza virus separately to examine such potential differences and the impact on influenza epidemic dynamics at the population level.

Our study also sidestepped several key factors that may shape influenza transmission dynamics, in particular, age, vaccination, and seasonal changes in contact patterns. Due to a lack of age-specific incidence data, we did not include age structure in our models. Vaccination rate in Hong Kong has increased in recent years, particularly among children aged <12 years and adults 65 years or older, due to vaccination subsidy provided to these age groups from 2008 onwards [42-44]. For instance, vaccination rate among children aged <12 years increased from 9.7% in 2011/12 to 55.4% in 2018/19; and vaccination rate among adults 65+ increased from 31.7% in 2011/12 to 43.6% in 2018/19 [45, 46]. These increases in vaccination coverage may in part explain the apparent decreases in ILI+ in recent years; a one-sided *t*-test indicated that the yearly ILI+ during 2011-2018 were significantly lower than years before 2009 (i.e., excluding the 2009 pandemic period; p=0.009). However, we do not have detailed data on vaccination rate nor vaccine efficacy over the entire study period in order to account for the impact of vaccination. Further, seasonal changes in contact patterns could also contribute to the observed seasonal influenza epidemic dynamics in Hong Kong. For instance, during a hot, humid summer, people may spend more time indoors and thus create additional opportunities for onward transmission to occur. Similar increases of indoor contact and transmission could occur during colder days in the winter. As such, the increased transmission under highly humid conditions in the summer or colder-drier conditions in the winter may be in part due to increased contact indoors, in addition to the higher survival rate of influenza viruses under these environmental conditions as shown in laboratory studies [15, 16, 39]. Our modeling here is not able to tease this apart. Future work is warranted to incorporate these additional factors that may further improve understanding of influenza transmission dynamics in tropical and subtropical climates. Nevertheless, we note that, all climate forcing models tested here are subject to the same limitations discussed above. Thus, the relative performance of the models and our general findings regarding key climate drivers still hold.

In summary, we have developed a simple mechanistic model incorporating the impact of AH and temperature on influenza transmission that is able to recreate the long-term influenza epidemic dynamics in Hong Kong, a subtropical city with highly diverse epidemic patterns. Past work has demonstrated that, incorporating information on AH into mechanistic influenza models significantly improved model fit and forecast accuracy in temperate regions [11]. Given that forecasts in the tropics and subtropics tend to be substantially less accurate than forecasts in temperate regions [36], the climate forcing model developed here could support a better understanding of climatic drivers of influenza transmission in these regions. Future work could incorporate this model into a SIRS model-forecast system to improve forecast accuracy in the tropics and subtropics to aid public health and medical workers in better anticipating influenza transmission in forthcoming weeks.

## Data Availability

Compiled daily temperature and specific humidity data along with model code are available at https://github.com/wan-yang/flu-subtropic-climate-models.   Influenza data (after 2014) are available from https://www.chp.gov.hk/en/statistics/data/10/641/642/2274.html

## Acknowledgements

We thank the Hong Kong Center for Health Protection for providing data on sentinel surveillance and laboratory surveillance for influenza. We also thank Columbia University Mailman School of Public Health for access to high performance computing.

## Supporting Information

### 1. Preliminary data processing

#### 1.1 Calculation of absolute humidity [1]

First, we calculated saturation vapor pressure as:

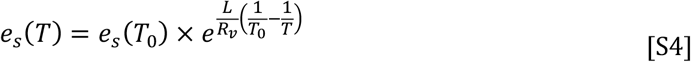

where *e*_*s*_*(T)* is the saturation vapor pressure at temperature *T* (in K), *e*_*s*_*(T*_*0*_) is the saturation vapor pressure at 273.15 K, *L* is the latent heat of evaporation for water, *R*_*v*_ is the gas constant for water vapor. We then computed vapor pressure at each time point as:

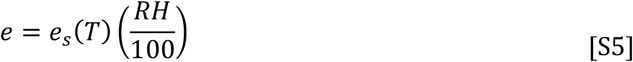

where *RH* represents relative humidity. We calculated the mixing ratio, *mr*, as:

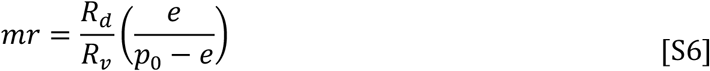

where *R*_*d*_ is the gas constant for dry air and *p*_*0*_ is the atmospheric pressure at sea level. Finally, absolute humidity was calculated as:

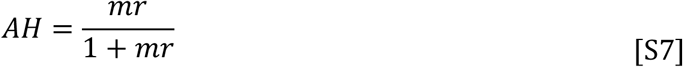

### 2. Simulation methods and modeling details

#### 2.1 Stochastic Model Runs

Similar to the climate forcing model used to model influenza in temperate region by Shaman et al. [2], we constructed a stochastic Markov chain, where stochastic is introduced by the transition between states. The number of individuals moving from one state to another (i.e. susceptible to infected, infected to recovered) is random draw from a Poisson distribution with a rate determined by Eqn 1.

#### 2.2 Climate forcing model for temperate regions (Null2)

The climate forcing model for temperate regions, first introduced by Shaman et al [2], assumes influenza transmission decreases monotonically with increasing absolute humidity. This relationship is modeled as:

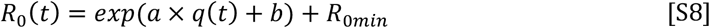

Where *a* = −180, *b*= log(*R*_*0max*_− *R*_*0min*_), *R*_*0min*_is the maximum daily basic reproductive number, while is the minimum daily basic reproductive number.

#### 2.3 Climate forcing model for (sub)tropical regions (AH/T, AH/T/Short, AH/T/Vary, AH/T/Strain)

Absolute humidity alone was proposed to have a bimodal effect on influenza transmission. And the formula can be written as follow:

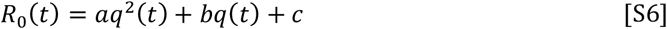

Values of *a, b*, and *c* are defined as:

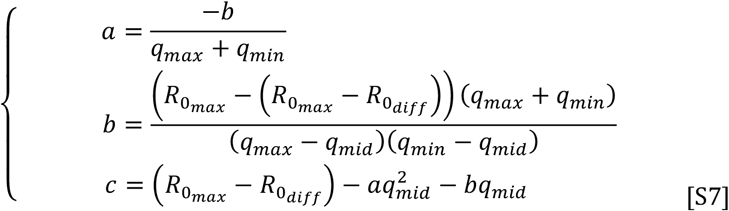

Given equation S6, the derivation of equation S7 is shown as below:

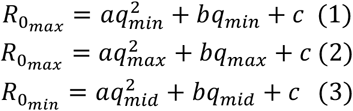

Subtracting (1) from (2), we can get:

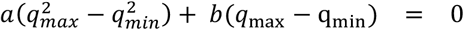

Rearrange the equation:

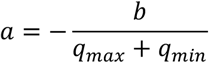

Moreover, by subtracting (3) from (2), we can get:

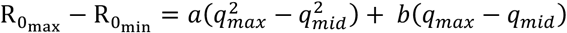

Plug in *a* :

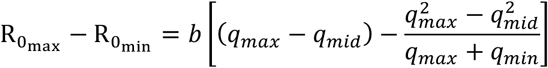

Move *b* to the left-hand side and rearrange:

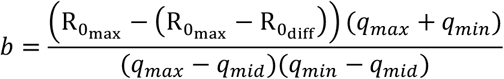

Plug *a* and *b* into Eqn. S7:

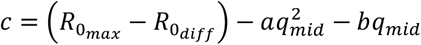

## Supplementary figures

**Fig S1.**
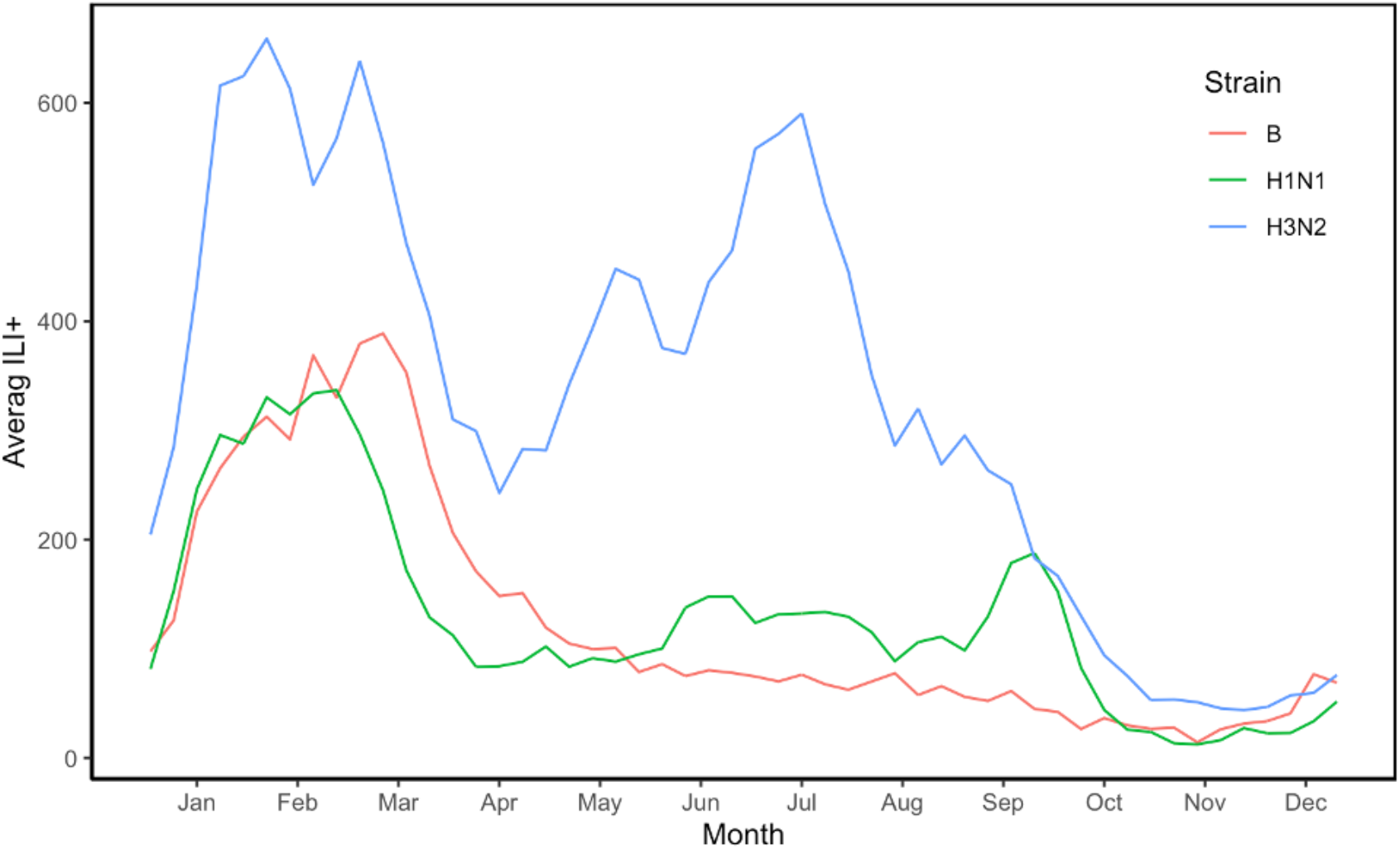
The change of mean ILI+ of each influenza (sub)type in circulation within a year. The mean ILI+ took average of the ILI+ observed in Hong Kong over 21 years (1998-2018).

**Fig S2.**
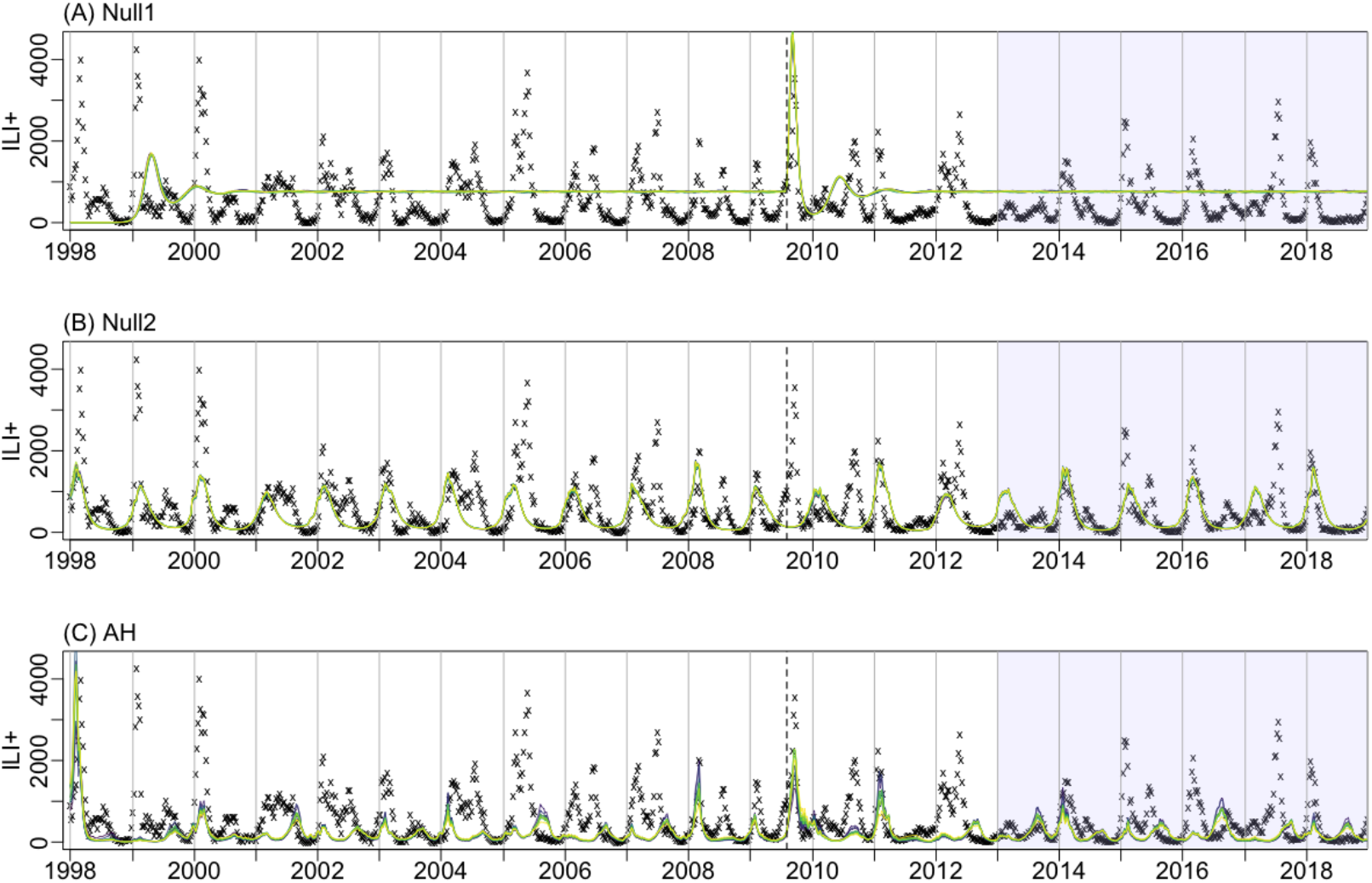
Top10 model fits for Null1 (A), Null2 (B), and AH (C) model. Black crosses show observed ILI+; the colored lines run through the crosses show the top 10 model estimates. The vertical dash line indicates the onset of the 2009 pandemic. The shaded regions indicate testing years (2013-2018); and the rest are the training years.

**Fig S3.**
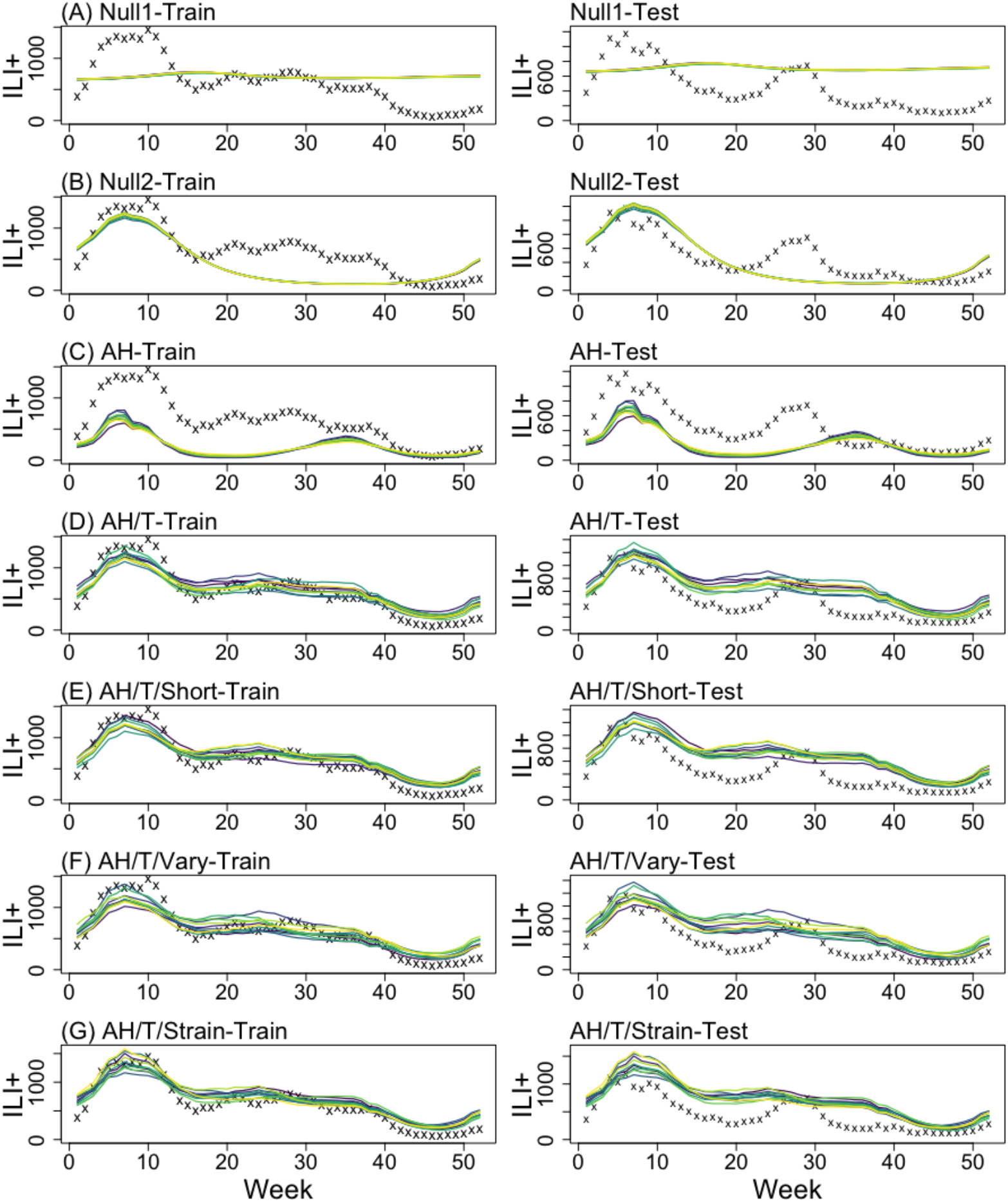
Top10 model fits for the observed seasonality (averaged over training or testing years) for the seven models: Null1 (A), Null2 (B), AH (C), AH/T (D), AH/T/Short(E) AH/T/Vary (F) and AH/T/Strain (G). Black crosses show observed averaged ILI+ over training or testing years; the colored lines run through the crosses are the top10 model estimates. Left panels are the model fit towards training data, and the right panels plot the fit for testing.

## Tables

**Table S1.**
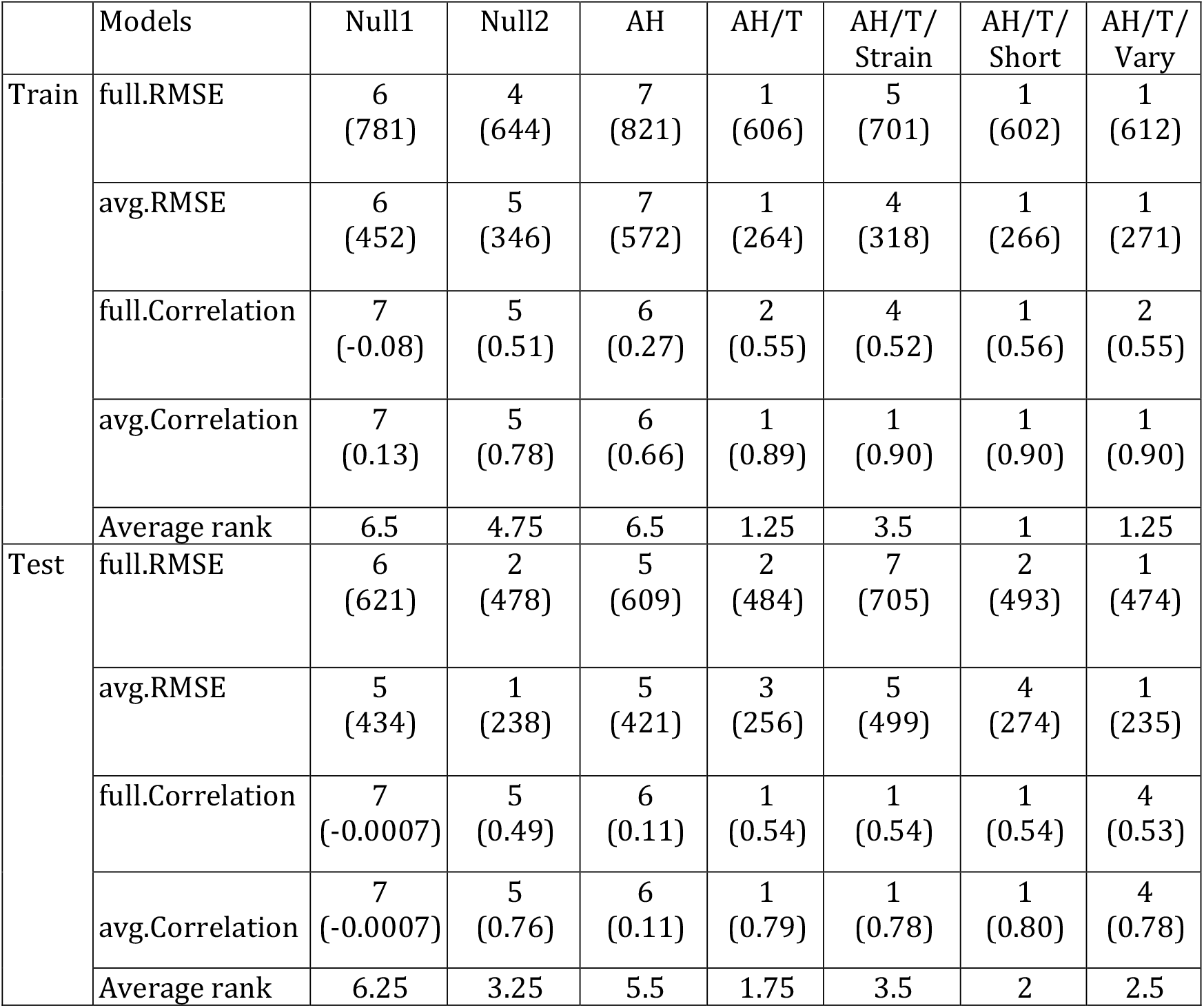
The model performance ranks for each comparison metric using training and testing data (excluding the 2009 pandemic). The rankings are determined by the model’s absolute mean rank differences with the best-ranked model. Different rankings between models indicate significantly different absolute mean ranks (i.e., p-value<0.007). The mean value of the corresponding metric is shown in the parenthesis.

|  | Models | Null1 | Null2 | AH | AH/T | AH/T/<br>Strain | AH/T/<br>Short | AH/T/<br>Vary |
| --- | --- | --- | --- | --- | --- | --- | --- | --- |
| Train | full.RMSE | 6<br>(781) | 4<br>(644) | 7<br>(821) | 1<br>(606) | 5<br>(701) | 1<br>(602) | 1<br>(612) |
|  | avg.RMSE | 6<br>(452) | 5<br>(346) | 7<br>(572) | 1<br>(264) | 4<br>(318) | 1<br>(266) | 1<br>(271) |
|  | full.Correlation | 7<br>(-0.08) | 5<br>(0.51) | 6<br>(0.27) | 2<br>(0.55) | 4<br>(0.52) | 1<br>(0.56) | 2<br>(0.55) |
|  | avg.Correlation | 7<br>(0.13) | 5<br>(0.78) | 6<br>(0.66) | 1<br>(0.89) | 1<br>(0.90) | 1<br>(0.90) | 1<br>(0.90) |
|  | Average rank | 6.5 | 4.75 | 6.5 | 1.25 | 3.5 | 1 | 1.25 |
| Test | full.RMSE | 6<br>(621) | 2<br>(478) | 5<br>(609) | 2<br>(484) | 7<br>(705) | 2<br>(493) | 1<br>(474) |
|  | avg.RMSE | 5<br>(434) | 1<br>(238) | 5<br>(421) | 3<br>(256) | 5<br>(499) | 4<br>(274) | 1<br>(235) |
|  | full.Correlation | 7<br>(-0.0007) | 5<br>(0.49) | 6<br>(0.11) | 1<br>(0.54) | 1<br>(0.54) | 1<br>(0.54) | 4<br>(0.53) |
|  | avg.Correlation | 7<br>(-0.0007) | 5<br>(0.76) | 6<br>(0.11) | 1<br>(0.79) | 1<br>(0.78) | 1<br>(0.80) | 4<br>(0.78) |
|  | Average rank | 6.25 | 3.25 | 5.5 | 1.75 | 3.5 | 2 | 2.5 |

